# Sexually transmitted infection knowledge levels, socio-demographic characteristics and sexual behaviour among men who have sex with men: results from a cross-sectional survey in Nairobi, Kenya

**DOI:** 10.1101/2023.02.02.23285368

**Authors:** Delvin Kwamboka Nyasani, Meshack Onyambu, Laura Lusike Lunani, Geoffrey Ombati Oino, Gaundensia Nzembi Mutua, Matt Price, Justus.O. Osero

**Affiliations:** Kenyatta University Department of Community Health and Epidemiology, Nairobi, Kenya; KAVI Institute Of Clinical Research, University Of Nairobi, Nairobi, Kenya; IAVI, New York, United States of America; Department of Epidemiology and Biostatistics, University of California, San Francisco, California, United States of America

## Abstract

**Background:** High rates of sexually transmitted infections (STIs) among men who have sex with men (MSM) have been reported, there is little research on their STI knowledge. Our study sought to characterize knowledge and awareness of STIs among MSM in Nairobi, Kenya.

**Methods:** We mobilized MSM aged ≥18 years from Nairobi into a cross-sectional study. To determine their understanding about STIs, a pre-tested structured questionnaire was administered. Knowledge score was generated by summing up the number of responses answered correctly by a participant. We dichotomized scores as “low” and “high”, by splitting the group at <12 and ≥12 which was the mean.

**Results:** A total of 404 participants were interviewed between the month of March and August 2020. The mean age was 25.2 (SD=6.4) years. Majority were single (80.4%) and Christians (84.2%). All participants had some formal education ranging from primary to tertiary; the majority (92.3%) had secondary education or more. Most (64.0%) were employed and their monthly income ranged from <50->150 USD. Almost all (98.5%) were Kenyans.

Of the 404 (90.6%) self-identified as male and (47.5%) reported to be exclusively top partners. Many (39.9%) reported being both bottom and top, while those reporting to be bottom partners were, (12.6%). The last 12 months, (55.4%) of the participants reported having sex with men only and (88.6%) reported to have had more than one sexual partner.

Participants scored an average of 12.2, out of 29 SD 4.5. The multivariable modelling revealed that participants aged ≥25 years were more likely to have a higher knowledge score compared with the participants aged 18-24 years (aOR=0.973, CI: 0.616-1.538). Regarding education and occupation, participants who had tertiary education and those who were employed were more likely to have a higher knowledge score compared with the participants who had primary education and were not employed (aOR=2.627, CI:1.142-6.043) (aOR=0.922, CI:0.401-2.117). Participants who were earning (USD >150) were more likely to have a higher knowledge score compared to the ones who were not earning (aOR=2.520, CI: 0.900-7.055). Further bisexual men were more likely to have a higher knowledge score compared with the participants who were having sex with men only (aOR= 1.550, CI: 1026-2.342)

**Conclusion:** Participant’s knowledge level regarding STIs was low. We recommend health care workers to continue educating patients about STIs.

## Introduction

### Background

Over 1 million sexually transmitted infections (STIs) are acquired daily globally; common among them are chlamydia, gonorrhoea, syphilis and trichomoniasis. Many of these infections are frequently mild or asymptomatic (1) In low- and middle-income countries (LMICs), symptomatic STIs are often treated by syndromic management which frequently misses asymptomatic STIs that therefore remain untreated ad potentially infectious (2). Regardless of symptoms, STIs can cause serious morbidity including cancer, infertility, and enhanced HIV transmission (3,4).

Prevalence rates of STIs vary from area to area, nation to nation and between urban and rural populations. The infections are more prevalent in Asia and sub-Saharan Africa (SSA), but SSA is estimated to contribute a greater proportion of the cases of STIs every year, highlighting a need for intervention (5).

The impact of STI epidemics is of particular importance among key, higher-risk populations who include: young people and adolescents, men who have sex with men (MSM), people in prisons, sex workers and people who inject drugs. These populations are often vulnerable to both HIV and other STIs not only because of their high-risk sexual and drug use behaviours, but also by structural barriers such as low access to quality health services, stigma and discrimination(6).

The risk may even be even greater among MSM because unprotected receptive anal sex; is a greater risk factor for STI and HIV acquisition than unprotected vaginal sex (7).

In developed countries such as United States, England and Netherlands the data as also shown that MSM bear a much higher burden of new cases of STI and HIV than their heterosexual male counterparts (8–11)

The situation is similar in Africa, where increasing attention has been spent describing the epidemiology of STIs and HIV among MSM in recent years. In Uganda, a study conducted among 300 MSM, found that they had a higher HIV prevalence 13.7% compared to the general adult male population at 4.5% (12).

In Kenya, a study conducted in Nairobi among 563 male sex workers and other MSM found that the prevalence of testing positive for one or more STIs combined (Syphilis, gonorrhoea or chlamydia) among male sex workers was higher (15.0%) compared to those who were not sex workers (5.3%) (13). Also according to (14) among the estimated 22,000 MSM in Kenya, the HIV prevalence was 18.2% compared to the national general population at 5.9%.

According to (15) comprehensive HIV and AIDS knowledge among sexually active men had reduced the odds of them contracting STIs. Being that the HIV prevalence among MSM in Kenya is estimated to be three times higher than that of the general population, we sought to characterize knowledge and awareness of STIs among MSM in Nairobi City County, Kenya.

## Materials and Methods

### Study design

This was a cross-sectional study among sexually active MSM within Nairobi, Kenya, conducted between the months of March-August 2020. We interviewed participants to determine their understanding of STIs and assessed for association with socio-demographic characteristic and sexual behaviour.

### Study participants

The study was designed to enrol men aged ≥18 years and who have sex with men from places where MSM are known to either solicit their clients or where sexual activity was common including “ cruising” areas along streets, or bars with or without lodging (16). They had to be willing and able to give informed consent to participate. MSM who were obviously intoxicated were excluded from participation.

### Participant recruitment

Mobilization was done by MSM community mobilizers. We identified MSM community mobilizers from the Nairobi-based Sex Workers Outreach Programme (SWOP) with whom we had partnered for a recent study (17). The community mobilizers gave a brief description of the research activities to potential participants. Then using a systematic random sampling procedure they selected participants with the desired characteristics. A systematic random sampling is a sampling technique that the researcher selects participants to be included in the sample based on a systematic rule, using a fixed interval.

We had twenty one days to enrol 422 participants. We had purposively selected 5 sub-counties that were densely populated with MSM(16). To get the percentage of the participants to be accessed from each sub-county, the researcher took the expected number in a specific sub-county divided by the total participants from all the 5 sub-counties and multiplied by 100.Then to get the numbers to be recruited from each sub county, it was the percentage of the participants to be accessed from each sub-county multiplied by the required number of participants (n=422) (Table 1).

**Table 1:** The Number of MSM in the selected Sub-Counties, at selected MSM geographic areas In Nairobi, Kenya and the sampled population.

| Sub-County | Estimated number of MSM available on all selected MSM geographic areas | Percentage of the total MSM to be accessed | Number of MSM required for the study ((n=422) |
| --- | --- | --- | --- |
| **Starehe** | 2000 (40 MSM geographic areas) | 55% | 232 |
| **Westlands** | 1000 (27 MSM geographic areas ) | 27% | 114 |
| **Dagoretti South** | 360 (20 MSM geographic areas) | 10% | 42 |
| **Ruaraka** | 180 (15 MSM geographic areas) | 5% | 21 |
| **Kasarani** | 100 (10 MSM geographic areas) | 3% | 13 |
| **Total** | 3640 | 100 | 422 |

To get the first participant from each sub-county on each day of mobilization, we had 2 sealed papers written “Yes and No”. The mobilizers issued the first two participants the sealed papers if for example, the second participant selected “Yes”, then they were issued with a unique identification card. Then they continued mobilizing the second participant until the required number was attained. If the sampled participant had declined to participate, the next exiting participant was sampled. The sampled participants were then referred to the researcher and the research assistants for interviews at a selected venue. The selected venue was a safe and private place identified by the mobilizers where the interviews could be conducted; without attracting attention to the participants. At the entrance to the selected venue, a peer navigator confirmed participant identity by confirming that they had a unique identification card and that they were not intoxicated. Thereafter informed consent was obtained from the participants before the questionnaire was administered. Mobilization from the MSM geographic areas was done on 4 selected peak days (i.e., Wednesday, Friday, Saturday and Sunday) as identified by the key population implementing partners during the key population size estimate mapping(16). During that time also the peak time identified was 18.00hrs-22.00hrs and that is what we adopted. Peak day refers to a day when the number of MSM present at the MSM geographic areas was more than usual.

### Procedure

Participants were assigned a unique study identification number. Data was collected about socio-demographics and sexual behaviour including: age, marital status, religion, education level, occupation, level of income, nationality, gender identity, roles during sexual activity, sex with whom in the past 12 months, number of sexual partners in the past 12 months, type of sex, if received gifts/money in exchange for sex, whether gave gifts/money in exchange for sex and history of having ever contracted any STI.

To determine their understanding about STIs participants were then given a pre-tested structured questionnaire (Supplemental material). The questionnaire had four open ended questions and one short answer question. Interviewers were instructed not to prompt participants but only tally all correct responses provided. If a participant gave an incorrect answer it was ignored but if it was correct and not among the options given it was captured as other and later analysed. The lead researcher in the field did quality control checks on the questionnaires before the participant was reimbursed, allowing the team to avoid missing data or clarify ambiguities.

### Study Variables

#### Dependent variable

Knowledge score was the dependent variable. In assessing knowledge levels the participants were asked the following questions; which diseases are spread via sexual intercourse? What the signs and symptoms of STIs were? How one could be infected with STIs? Can someone be infected with STIs via anal sexual intercourse? And Through which media could one get infected with STIs? (Additional details are shown in the supplemental material). Interviewers were instructed not to prompt participants but only tally all correct responses provided.

A knowledge score was generated by summing up the number of responses answered correctly by a participant; to give a score ranging from 0 to 29. For the purposes of analysis, we dichotomized scores as “low” and “high”, by splitting the group at <12 and ≥12 (additional details are shown in Appendix 1).

### Independent variables

Independent variables included in this analysis included: age, marital status(single never married, married and single ever married), religion (Christians and non-Christians), education (primary, secondary and post-secondary), occupation (unemployed and employed), level of income (No income, <5000, 5001-10,000, 10001-15000 and >15000), nationality (Kenyans and non-Kenyans), gender identity (male, transgender woman, intersex and non-conforming), roles during sexual activity (top, bottom or versatile), sex with whom in the past 12 months (men or both men and women), number of sexual partners in the past 12 months (one or more than one), type of sex (vaginal, anal, oral, mutual masturbation), received gifts/money in exchange for sex (yes, all the time, yes, sometimes, No), gave gifts/money in exchange for sex (yes, all the time, yes, sometimes, No), and ever contracted STI (Yes, No) (see Table 1 and 2).

**Table 2:** Socio-demographic characteristics of study participants and their association with participant’s knowledge score.

| Characteristics | Total<br>(n=404) | Knowledge score |  | P Value |
| --- | --- | --- | --- | --- |
|  |  | <12<br>(n=215) | ≥12<br>(n=189) |  |
|  | n(%) | n(%) | n(%) |  |
| Mean age=25.20, SD=6.41 |  |  |  | 0.129 |
| Age bracket |  |  |  |  |
| 18-24 | 241(59.7) | 127(52.7) | 114(47.3) |  |
| ≥25 | 163(40.3) | 73(44.8) | 90(55.2) |  |
| Marital Status |  |  |  | 0.859 |
| Single never married | 325(80.4) | 162(49.8) | 163(50.2) |  |
| Married | 54(13.4) | 27(50.0) | 27(50.0) |  |
| Single ever married | 25(6.2) | 11(44.0) | 14(56.0) |  |
| Religion |  |  |  | 0.135 |
| Christian(Catholic & protestants) | 340(84.2) | 174(51.2) | 166(48.8) |  |
| Non-Christian (Muslim, Atheists, Hindu) | 64(15.8) | 26(40.6) | 38(59.4) |  |
| Education level |  |  |  | 0.001 |
| Primary | 31(7.7) | 19(61.3) | 12(38.7) |  |
| Secondary | 226(55.9) | 126(55.8) | 100(44.2) |  |
| Post-secondary | 147(36.4) | 55(37.4) | 92(62.6) |  |
| **Occupation** |  |  |  | 0.007 |
| Unemployment/Student | 145(36.0) | 85(58.6) | 60(41.4) |  |
| Employment/Self-employed | 259(64.0) | 115(44.4) | 144(55.6) |  |
| **Level of income** |  |  |  | 0.005 |
| None | 120(29.7) | 75(62.5) | 45(37.5) |  |
| <5000 | 54(13.4) | 28(51.9) | 26(48.1) |  |
| 5001-10000 | 87(21.5) | 38( 43.7) | 49(56.3) |  |
| 10001-15000 | 71(17.6) | 33(46.5) | 38(53.5) |  |
| >15000 | 72(17.8) | 26(36.1) | 46(63.9) |  |
| **Nationality** |  |  |  | 0.446 |
| Kenyans | 398(98.5) | 196(49.2) | 202(50.8) |  |
| Non-Kenyans | 6(1.5) | 4(66.7) | 2(33.3) |  |
| **\* KEY: by Chi-square test or Fisher’s Exact Test where cell size was small** |  |  |  |  |

### Data analysis

Data was entered into statistical package for social sciences programme (IBM-SPSS) version 25. Quantitative data from the study questionnaires was coded. Single data entry was done into SPPS database after quality control checks had been done. Further cleaning was carried out after data entry using frequency distributions and cross tabulations until no more errors were detected. Participant’s characteristics are presented by use of frequencies and percentages for categorical variables. Pearson’s Chi-square and Fishers exact test where appropriate was done to characterize any statistical significant associations between STI knowledge, demographics and reported sexual behaviour. Variables with a *p*≤0.05 and age being a priori variable were included in a multivariable logistic regression model to characterize factors associated with higher or lower participants STI knowledge.

### Ethical consideration and study approval number

Approval of the study was sought from Kenyatta University Board of Post Graduate studies. Ethical Clearance was given by Kenyatta University ethics review committee; (PKU/1071/11121). Permission to conduct the study was sought from the National Council for Science, Technology and Innovations (NACOSTI), and from Nairobi County and sub-county facilities. Confidentiality and anonymity of the information given by the participants was protected by ensuring that the names of the participants were not indicated in the data collection tools. Participants were reimbursed in the local currency equivalent of $2 USD in compensation for their time.

## Results

### Demographics

A total of 404 participants were interviewed between the month of March and August 2020. They had a mean age of 25.2 (SD=6.4) years. The majority were single never married (80.4%; 325) and Christians (84.2%; 340). All participants had some formal education ranging from primary to tertiary level; the majority (92.3%; 373) had secondary education or more. Most of the participants (64.0%; 259) were employed and their level of monthly income ranged from $<50->150 USD. Almost all the participants (98.5%; 398) were Kenyans. (Table 2)

#### Sexual behaviour

Most of the participants (90.6%; 366) self-identified as male and almost half (47.5%; 192) of them reported to be exclusively insertive (“Top”) partners. Many (39.9%; 161) reported being versatile (both bottom and top), while those reporting to be receptive (“Bottom”) partners were, (12.6%; 51). In the last 12 months, (55.4%; 224) of the participants reported having sex with men only and (44.6 %; 180) reported being bisexual. Also during that period, majority of the participants (88.6%; 358) reported to have had more than one sexual partner (Table 2).

Majority of the respondents (93.6%; 378) reported to have had anal sex, (42.1%; 170) had vaginal sex, (28.5%; 115) reported oral and (19.6%; 79) reported mutual masturbation. Almost half (47.5%; 192) of the participants, reported receiving gifts/money in exchange for sex, sometimes. Majority of the participants (95.3%; 385), engaged in transactional sex. Participants who reported to have ever contracted STI were (43.3%; 175). (Table 3)

**Table 3:**
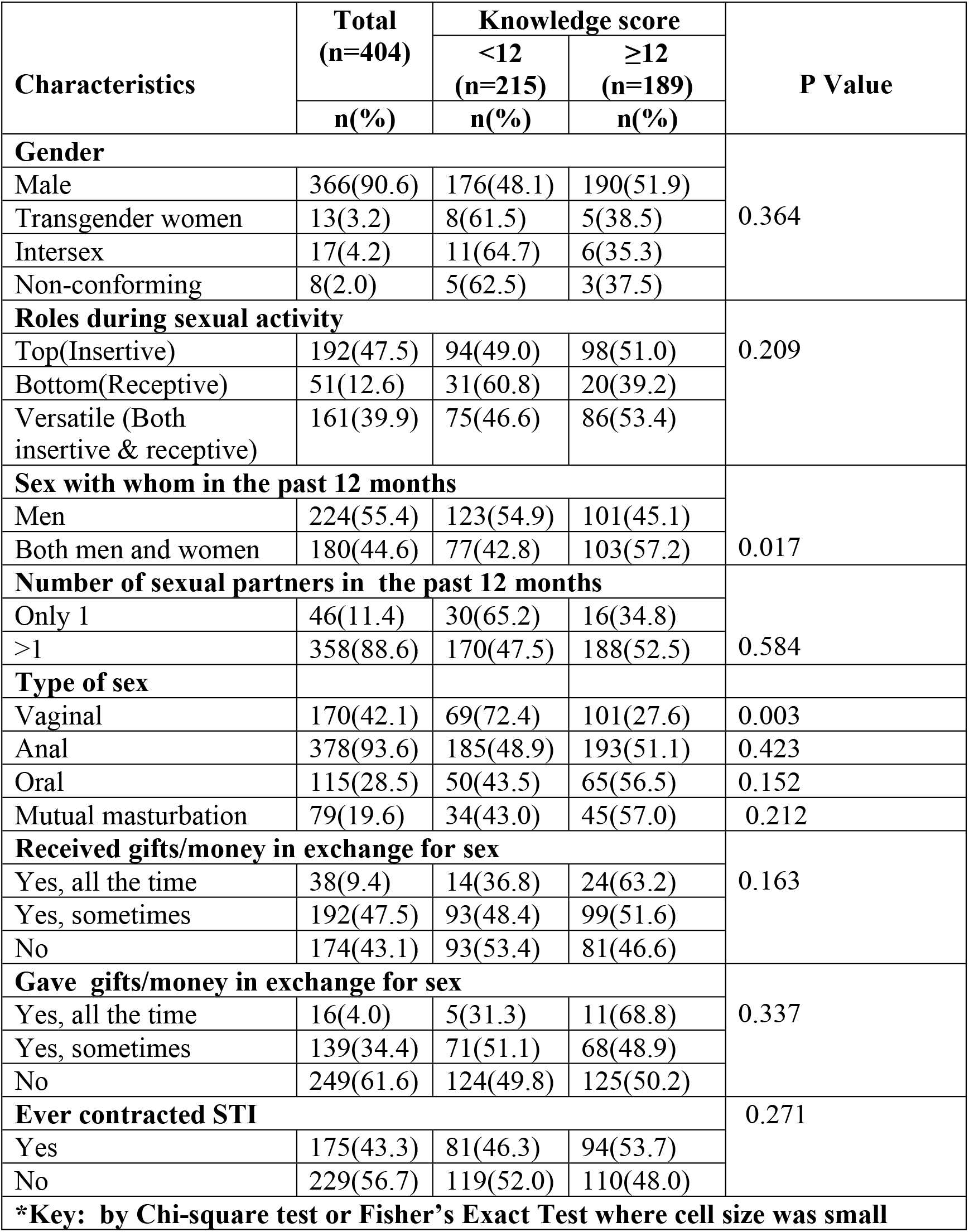
Sexual behavioural characteristics of study participants and their association with participant’s knowledge score.

#### STI knowledge

Out of a total of 29 possible correct answers the highest participant scored 25, we observed an average score of 12.2 correct, SD 4.5 (additional details shown in Appendix 1).As indicated on Table 4 below, majority of the participants were aware of gonorrhoea (92.8 %; 375), syphilis (88.1%; 356), HIV (65.6%; 265), genital warts (33.9%; 137), Herpes Simplex (22.8%; 92), chancroid (20.5%;83), hepatitis B (17.3%;70), chlamydia (13.6%;55) and trichomoniasis (5.2%;21).

**Table 4:**
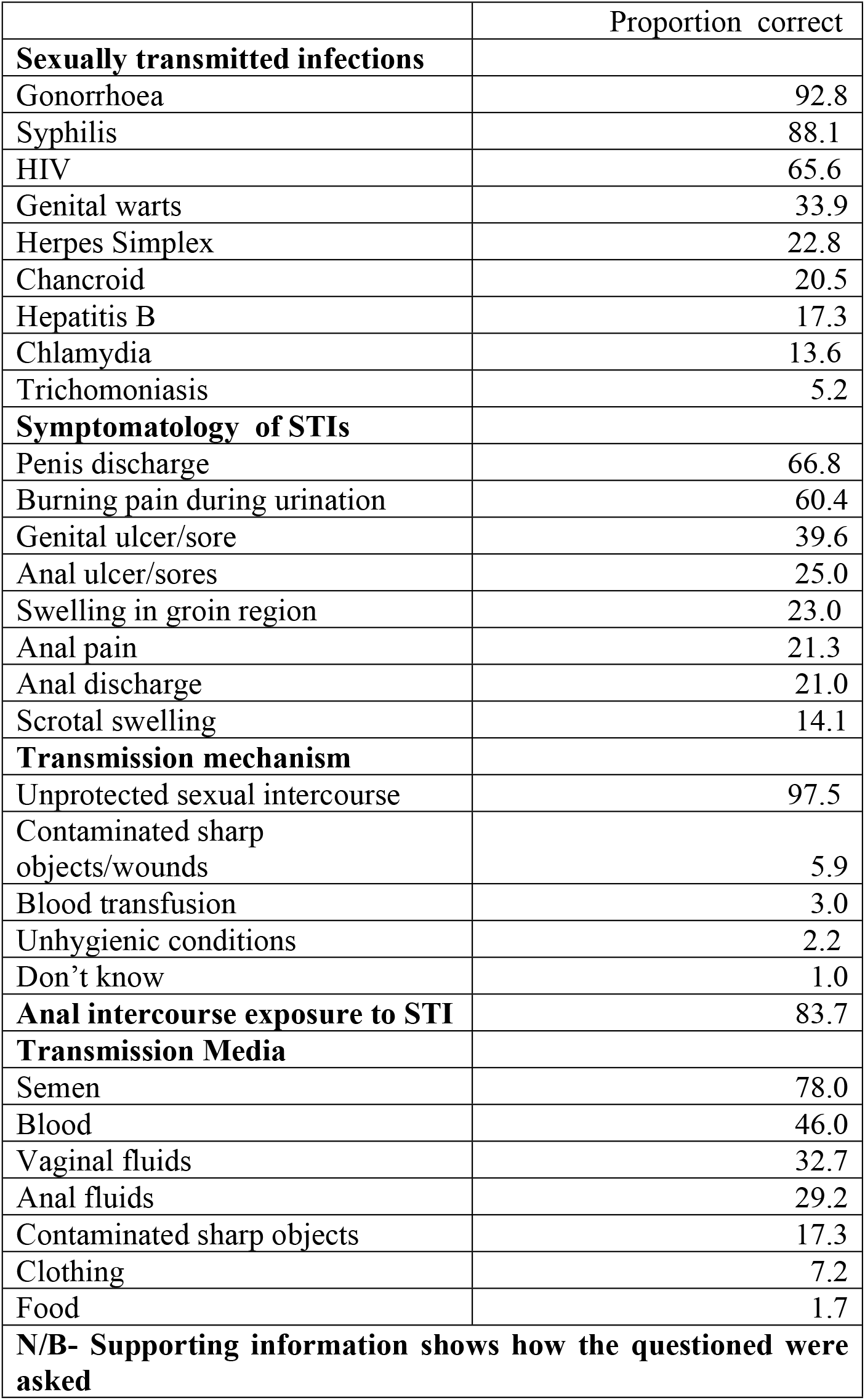
Proportion of participants answering STI knowledge items correctly.

|  | Proportion correct |
| --- | --- |
| **Sexually transmitted infections** |  |
| Gonorrhoea | 92.8 |
| Syphilis | 88.1 |
| HIV | 65.6 |
| Genital warts | 33.9 |
| Herpes Simplex | 22.8 |
| Chancroid | 20.5 |
| Hepatitis B | 17.3 |
| Chlamydia | 13.6 |
| Trichomoniasis | 5.2 |
| **Symptomatology of STIs** |  |
| Penis discharge | 66.8 |
| Burning pain during urination | 60.4 |
| Genital ulcer/sore | 39.6 |
| Anal ulcer/sores | 25.0 |
| Swelling in groin region | 23.0 |
| Anal pain | 21.3 |
| Anal discharge | 21.0 |
| Scrotal swelling | 14.1 |
| **Transmission mechanism** |  |
| Unprotected sexual intercourse | 97.5 |
| Contaminated sharp objects/wounds | 5.9 |
| Blood transfusion | 3.0 |
| Unhygienic conditions | 2.2 |
| Don't know | 1.0 |
| **Anal intercourse exposure to STI** | 83.7 |
| **Transmission Media** |  |
| Semen | 78.0 |
| Blood | 46.0 |
| Vaginal fluids | 32.7 |
| Anal fluids | 29.2 |
| Contaminated sharp objects | 17.3 |
| Clothing | 7.2 |
| Food | 1.7 |
| **N/B- Supporting information shows how the questioned were asked** |  |

Regarding signs and symptoms of STIs, most of the participants were cognizant of penile discharge (66.8%; 270), burning sensation during urination (60.4%; 244), genital ulcer/sores (39.6%; 160), anal discharge (21.0%; 85), swelling in the groin region (23.0%; 93), anal ulcer/sore (25.0 %; 101), anal pain (21.3%; 86), and scrotal swelling (14.1%; 57).

Also almost all the participants knew that STIs were transmitted through unprotected sexual intercourse (97.5%; 394) i.e. vaginal, anal, oral and mutual masturbation. Also about the anal sexual intercourse risk to STIs, majority of the participants (83.7%; 338) were aware that they could be infected with STIs through unprotected anal sexual intercourse. About three quarters of the participants reported correctly that they could be infected with STIs through semen (78.0%; 315) and through blood (46.0%; 186), vaginal fluids (32.7%; 132), contaminated sharp objects (17.3%; 70) and anal fluid (29.2%; 118).

#### Participant characteristics independently associated with participant’s knowledge about STIs

The multivariable modelling revealed that participants who were aged ≥25 years were more likely to have a higher knowledge score compared with the participants who were aged 18-24 years (adjusted odds ratio aOR=0.973, 95% CI 0.616-1.538). Regarding education, participants who had tertiary education were three times more likely to have a higher knowledge score compared with the participants who had primary education (aOR=2.627, 95% CI 1.142-6.043). Regarding occupation participants who were employed had a higher knowledge score compared to the ones who were not employed (aOR=0.922, 95% CI 0.401-2.117). Under level of income, participants earning Kshs 5000-10,000 were three times likely to have a higher knowledge score compared to the ones who were not earning (aOR 2.332, 95% CI 0.990-6.263). Participants who were earning Kshs >15000 (USD >150) were also three times more likely to have a higher knowledge score compared to the ones who were not earning (aOR=2.520, 95% CI 0.900-7.055). Further bisexual men were more likely to have a higher knowledge score compared with the participants who were having sex with men only (aOR= 1.550, 95% CI 1026-2.342) (Table 5).

**Table 5:** Participant characteristics independently associated with participant’s knowledge about STIs.

| **Variable** | **Unadjusted Odds Ratio (95% CI** | **P-Value** | **Adjusted Odds Ratio (95% CI** |
| --- | --- | --- | --- |
| **Age bracket Reference 18-24yrs** |  | 0.129 |  |
| >25 yrs | 1.373 (0.922-2.047) |  | 0.973 (0.616-1.538) |
| **Highest reported education level Reference- "Up to completed Primary"** |  | 0.001 |  |
| "Up to completed Secondary | 1.257 (0.582-2.711) |  | 1.284 (0.575-2.865) |
| "Up to completed Post-secondary | 2.648 (1.195- 5.872) |  | 2.627 (1.142-6.043) |
| **Occupation Reference (Not employed/student** |  | 0.007 |  |
| Employed | 1.774 (1.176-2.677) |  | 0.922 (0.401-2.117) |
| **Level of income Reference (No income).** |  | 0.005 |  |
| <5000 | 1.548 (0.809-2.962) |  | 1.857 (0.725-4.759) |
| 5000-10000 | 2.149 (1.225-3.771) |  | 2.491 (0.990-6.263) |
| 10001-15000 | 1.919 (1.059-3.480) |  | 1.940 (0.746-5.042) |
| >15000 | 2.949 (1.608-5.408) |  | 2.520 (0.900-7.055) |
| **Sex with whom Reference (Men)** |  | 0.017 |  |
| Bisexual | 1.629 (1.097-2.419) |  | 1.550 (1026-2.342) |

## Discussion

STI knowledge and awareness is a key element towards decreasing the incidence of STIs among populations at high risk of infection. However, findings from this study among MSM in Nairobi, Kenya, demonstrated low STI knowledge.

Participants in this study who reported HIV as one of the STIs were less compared to the one who reported Gonorrhoea and Syphilis. It’s expected that all the participants should be aware of it because HIV has no cure and it has caused devastating effects globally(18). This disconnect among some of the participants could be because patients on antiretroviral drugs look healthy. This has changed the perception of HIV/AIDS from a very fatal to a chronic and potentially manageable disease. Also the availability and administration of antiretroviral therapy has significantly reduced mortality and morbidity linked with HIV and AIDS(19). This disconnect of HIV as one of the STIs among some of the participants is worthy worrying about; because the progress that has been made to curb the spread of HIV might be futile. HCWs need to continue emphasizing to STI patients and other people in the society that HIV is one of the STIs and it has no cure. Being infected by other STIs such as Herpes Simplex Virus type 2 (HSV-2) and Syphilis infections puts them at a higher risk of being infected by HIV (3,4).

Others have found similar results; a study conducted among sixty racially –and ethnically diverse MSM and transgender women had also reported that the participants had low knowledge about STIs (20). Also another study conducted among 50 at risk MSM at Boston USA had found that the MSM lacked knowledge about the signs and symptoms of STDs (21). However, worth noting is that the measures of STI knowledge used on the above past studies varied greatly from those used in our survey. There is need for the participants to be health educated about STIs as some studies have revealed that patients who are enlightened have higher probability of seeking health care services compared to the once who are not (22). By them seeking treatment it can help stop the transmission of STI to others. To take care of the asymptomatic STIs; quarterly STI screening should be encouraged among MSM who are at risk of STIs.

In this study some of our participants reported of having oral sexual intercourse. Therefore, there is need for at least annual screening of extra-genital sites (rectum and pharynx) among sexually active MSM(23); so that, the extra-genital STIs cannot be missed out.

This study did not reveal statistical significant association between knowledge of the MSM and their age. Others have found similar results too; a study that was conducted among MSM in Ireland reported that participants who were aged 18-24 years of age had lower knowledge about STIs compared with the older MSM (24). Low knowledge about STIs among the youth could be occasioned by government policies and laws that criminalize key population behaviours and by education and health systems that pay no attention to or reject them(6).

Participants who had tertiary education were three times more likely to have a higher knowledge score compared with the participants who had primary education. Consistent with our findings is a study which was conducted in Melaka Malasyia; they found that participants who were studying in a degree program were two times more likely to be knowledgeable about STIs than the one who were not (25)

Further this study revealed that sexual behaviour of the study participants in terms of sex with whom, bisexual men had higher odds of knowledge about STIs compared to gay men. These findings are contrary to (26,27) who reported that bisexual men had limited availability of culturally sensitive education materials or health information that is specifically targeted to their needs. However, their settings are different from our Kenyan context.

Also majority of the participants had more than one sexual partner. Contrary to our findings, a study which was conducted among MSM in UK revealed that men who were knowledgeable about STI had more sexual partners. The authors of that study suggested that, in addition to knowledge, behaviours were determined by a complex range of psychological and eco-social factors (28). Still consistent with the UK study is a study which was conducted among 772 subjects in Estonia. They found that higher knowledge scores were not associated with lower HIV infection rates (29). This affirms that while knowledge is a key component of behaviour change models, by itself it is often insufficient to effect behaviour change. Models such as the Information-Behavioural Skills Model suggest that to take steps to reduce risk of STI infection, individuals would need not only knowledge about STIs, but also motivation to prevent it and the skills necessary to implement risk reduction actions, such as getting regularly tested for STIs (30).

### Limitations

Our study has several limitations. Our sample represents those men who seek sex at selected geographic areas in Nairobi, and may not be representative of MSM, but otherwise do not visit these areas. However, our aim was to enrol very high at risk individuals.

Our data are cross sectional, and as such we cannot infer causality between our independent variables and our dependent variable, knowledge of STIs.

### Implications to programmes

Individual health education and psychological approaches help an individual to acquire knowledge and skills to enact new behaviours to reduce risk (31). In this study participants had low knowledge about STIs and majority of them had more than one sexual partner. We do recommend HCWs to continue giving health talks about STIs to MSM during their health facility visits and in any other forums to enlighten them about STIs. Also because other studies have reported that knowledge alone is often insufficient to effect behaviour change. There is need for active follow-up of high risk individuals to prevent STIs which can be done by;

Recalling of MSM for follow-ups, who report engaging in high risk sexual behaviours via phone calls, peer leaders or community health works and creating demand for STI prevention cascade. Also bulky short messages reminder for offering STI testing to high risk MSM can be facilitated by using prompts incorporated within electronic MSM records.

On the other hand because there is a higher likelihood of extra-genital STIs being missed out by both the patient and clinicians; there is need for at least annual screening of extra-genital sites (rectum and pharynx) among sexually active MSM.

Also because asymptomatic STIs can be missed out due to syndromic management of STIs; we recommend quarterly STI screening for individuals who are at risk so that the asymptomatic STIs can be detected and treated to curb the spread of STIs.

### Implications to Policy Makers

Clear policies and guidelines should be put in place to enable HCWs to know what to do once an individual has self-identified himself as MSM. For instance, MSM can be linked to programmes or health facilities that can follow them up for possible regular screening and treatment of STIs.

Self-testing for STIs allows convenience, privacy and gives the patient an opportunity to avoid embarrassment while still accessing STI care. So they can implement the use of self-sampling kits for various STI testing as another study reported that HIV self –testing was accepted by similar populations (32).

## Conclusion

Participant’s knowledge level regarding STIs was low. Even though other studies have reported that individuals to reduce risk of STI infection would need not only knowledge about STIs. Still we recommend HCWs to continue educating patients about STIs. This is because patients who are enlightened have a higher probability of seeking health care services compared to the once who are not. Hence stop further spread of the STIs to their sexual partners.

## Data Availability

All relevant data are within the manuscript and its Supporting Information files.

## Supporting Information

### Supplemental questionnaire

indicating the questions assessing participant’s knowledge levels.

## Acknowledgements

This work was partially funded by IAVI with the generous support of USAID and other donors; a full list of IAVI donors is available at www.iavi.org. The contents of this manuscript are the responsibility of IAVI and co-authors and do not necessarily reflect the views of USAID or the US Government. We would like to acknowledge the participants and the research assistants who assisted in data collection. We also appreciate Kenneth Ekoru from the imperial university and Janet Muasya from the University of Nairobi for their input in the data analysis.

## Author Contributions

**Conceptualization:** Delvin Kwamboka Nyasani

**Data curation:** Delvin Kwamboka Nyasani and Laura Lunani Lusike

**Formal analysis:** Justus.O.Osero and Laura Lunani Lusike

**Funding acquisition:** Delvin Kwamboka Nyasani

**Project Administration:** Justus.O.Osero and Meshack Onyambu

## Resources

**Supervision:** Justus.O.Osero and Meshack Onyambu

**Validation:** Matthew A. Price and Gaundensia Nzembi Mutua

**Writing original draft:** Delvin Kwamboka Nyasani

**Writing, review & editing:** Delvin Kwamboka Nyasani, Justus.O.Osero, Meshack Onyambu, Geoffrey Oino Ombati, Matthew A. Price and Gaundensia Nzembi Mutua.

## Appendix 1: Knowledge assessment score of study participants

| **Knowledge Score** |  |  |  |
| --- | --- | --- | --- |
| **Mean=12.25,** |  | **Median=12.00** |  |
| **No. of right answers** | **No. of participants** | **Score percentage** | **Cumulative score percentage** |
| 3 | 1 | .2 | .2 |
| 4 | 5 | 1.2 | 1.5 |
| 5 | 5 | 1.2 | 2.7 |
| 6 | 16 | 3.9 | 6.7 |
| 7 | 24 | 5.9 | 12.6 |
| 8 | 30 | 7.4 | 20.0 |
| 9 | 36 | 8.9 | 28.8 |
| 10 | 40 | 9.9 | 38.7 |
| 11 | 44 | 10.8 | 49.5 |
| 12 | 33 | 8.1 | 57.6 |
| 13 | 32 | 7.9 | 65.5 |
| 14 | 46 | 11.3 | 76.8 |
| 15 | 15 | 3.7 | 80.5 |
| 16 | 15 | 3.7 | 84.2 |
| 17 | 12 | 3.0 | 87.2 |
| 18 | 14 | 3.4 | 90.6 |
| 19 | 8 | 2.0 | 92.6 |
| 20 | 5 | 1.2 | 93.8 |
| 21 | 3 | .7 | 94.6 |
| 22 | 5 | 1.2 | 95.8 |
| 23 | 5 | 1.2 | 97.0 |
| 24 | 4 | 1.0 | 98.0 |
| 25 | 8 | 2.0 | 100.0 |
| Total | 406 | 100.0 |  |

## Notes

### Competing Interest Statement

The authors have declared no competing interest.

### Funding Statement

DKN received a scholarship to study her Masters degree. Conception and design of the study was done by DKN.This work was partially funded by IAVI with the generous support of USAID and other donors a full list of IAVI donors is available at www.iavi.org. The funders had no role in study design, data collection and analysis, decision to publish, or preparation of the manuscript

### Author Declarations

Ethical consideration and study approval number Approval of the study was sought from Kenyatta University Board of Post Graduate studies. Ethical Clearance was given by Kenyatta University ethics review committee (PKU/1071/11121). Permission to conduct the study was sought from the National Council for Science, Technology and Innovations (NACOSTI), and from Nairobi County and sub-county facilities. Confidentiality and anonymity of the information given by the participants was protected by ensuring that the names of the participants were not indicated in the data collection tools. Participants were reimbursed in the local currency equivalent of $2 USD in compensation for their time.

